# If we cannot eliminate them, should we tame them? Mathematics underpinning the dose effect of virus infection and its application on covid-19 virulence evolution

**DOI:** 10.1101/2021.06.30.21259811

**Authors:** Zhaobin Xu, Hongmei Zhang

## Abstract

There is a dose effect in the infection process, that is, different initial virus invasion loads will lead to nonlinear changes in infection probability. Experiments already proved that there was a sigmoid functional relationship between virus infection probability and inoculum dose. By means of mathematical simulation of stochastic process, we theoretically demonstrate that there is a sigmoid function relationship between them. At the same time, our model found three factors that influence the severity of infection symptoms, those are virus toxicity, virus invasion dose and host immunity respectively. Therefore, the mortality rate cannot directly reflect the change of virus toxicity, but is the result of the comprehensive action of these three factors. Protective measures such as masks can effectively reduce the severity of infection while reducing the probability of infection. Based on the sigmoid function relationship between virus infection probability and initial virus invasion dose, we deduce that for highly infectious viruses, such as SARS-COV-2, the evolution of its toxicity is closely related to the host population density, and its toxicity will first increase and then decrease with the increase of host population density. That is to say, on the basis of extremely low host population density, increasing population density is beneficial to the development of virus towards strong toxicity. However, this trend is not sustainable, and there is a turning point of population density. Beyond this turning point, increasing population density will be beneficial to the development of virus towards weak toxicity. This theory can well explain the differences of mortality in Covid-19 in different countries. Countries with high population density and extremely low population density often correspond to lower mortality, while countries with population density in the range of 20-100/km^2^ often have higher mortality. At the same time, we propose that social distance and masks can effectively accelerate the evolution of virus towards low toxicity, so we should not give up simple and effective protection measures while emphasizing vaccination.

**Highlights:** Through mathematical simulation of random process, we prove the sigmoid function relationship between virus infection probability and initial virus invasion dose theoretically.

Our model found three factors that influence the severity of infection symptoms: virus toxicity, virus invasion dose and host immunity. This can help explain why the average infection age was declining as the epidemic went through.

With the increase of host population density, virus toxicity will increase at first and then decrease, which will explain the difference of mortality in different population density areas.

From the mathematical level, social distance, masks and other protective measures were proved to be positive in promoting the virus evolving into the less toxicity one. Vaccination could also promote virus virulence attenuation.

## Introduction

As of July 1, 2021, the COVID-19 epidemic has caused 180 million infections and more than 3.9 million deaths worldwide, becoming the largest public security crisis facing the world after World War II. The terrible thing about Covid-19 lies not only in its high transmission level, but also in its strong virulence, which displayed a much higher fatality rate compared with other common airborne viruses [1-3].

With the emergence of various virus variants and the existence of antibody self-attenuation, more and more evidences have gradually verified and are continually proving the failure of classic herd immunity theory in Covid-19 [4-6]. For example, the antibody positive rate in Manaus has reached 76% in October 2020, but there was still a large-scale outbreak in December 2020 [7]. In some areas of Iran, where natural immunization was adopted, and in August 2020, in some provinces with serious epidemic situation, such as Rasht city, the positive rate of antibody has exceeded the threshold of herd immunity [8]. These reports are not occasional. With the large-scale vaccination, this phenomenon reappeared in a larger geographical scope. For example, in countries with high vaccination coverage, such as Britain and Israel, the epidemic situation alleviated in short time due to vaccination, but this control is short-term and unsustainable. Because of the effects of virus mutation and antibody attenuation, the epidemic will resurge in a certain period in the future. Therefore, large-scale vaccination cannot eradicate the virus in a short time. The Bayesian model established by us pointed out the defects of herd immunity theory from the level of mathematical modeling, especially when we face of airborne RNA viruses with high mutation rate and strong transmission capacity [9].

In this case, a new viewpoint arises at the historic moment, people pay more attention to the change of virus toxicity [10]. We hope that Covid-19 will evolve toward the direction of low toxicity. If the toxicity of Covid-19 can be reduced to the level of influenza virus in the future, we can also get rid of epidemic in another way. We hope to find out the principle underpinning Covid-19’s virulence evolution, which will help to take appropriate interventions to influence the evolution direction and promote the virus to develop towards low toxicity. However, unfortunately, this kind of research is almost blank at present. Although the research on the evolution direction of virus virulence has a long history, in most cases, people only revealed the fact that virus toxicity varied and evolved, they have not found the driving factors of virus virulence evolution [11-13]. Our research aims to find out the relationship between the evolution direction of virus toxicity and population density, and promote the virus to develop towards low toxicity through artificial intervention, thus providing a new solution to overcome the tricky epidemic.

To study the relationship between virus toxicity evolution and host population density, we should first pay attention to the dose effect of virus infection, that is, the relationship between infection probability and initial invasion virus. Early models presumed that there was a linear relationship between them, which was obviously incorrect. Experiments in recent years have confirmed that there was a sigmoid function relationship between them [14-16]. Some researchers have verified this sigmoid relationship from the level of mathematical deduction [17]. The derivation seems elegant and reasonable at first glance, but there were lots of loose logics and mal-assumptions after a careful analysis. We will give a detailed discussion in the Results section. Different from the adaptation of ordinary differential equations, we established a stochastic modeling approach that could mimic virus proliferation in vivo, and deduced that there was indeed a sigmoidal relationship between virus dose and infection probability, not a linear or logistics one.

Based on this sigmoid relationship, we further studied the relationship between virus toxicity evolution and population density. Our model emphasizes the pressure of environment selection, not the mutation direction. We assume that the mutation of virus is random, and the evolution direction of virus toxicity does not depend on the mutation direction, but on the selection pressure from the environment. We use the reproductive coefficient R0 as the indicator of evolution, that is, the strain with larger R0 value will dominant evolution direction. By establishing a simple mathematical model using two viruses with different toxicity but same R0 value, we can calculate and simulate their R0 alternations under different host population densities to determine which strains will surpass the other one in long term evolution, and thus determine the influence of host population density on virus toxicity. Our results have been well verified in mortality in different countries, which can effectively explain many mysterious phenomena in the development of epidemic situation, and at the same time provide theoretical basis and guiding direction for artificial intervention to reduce virus toxicity.

## 1 Methods

### 1.1 Stimulation of virus in-vivo reproduction

The process of virus proliferation in vivo is a complicated process, but the overall virus load will first increase and then decrease. Our model has the following assumptions, which basically conform to the characteristics of virus proliferation in vivo.

#### Assumption 1

Single virus may die within a time interval, and the reason for death is that the virus has a half-life due to the influence of the surrounding environment. Antibody generated in host cell will shorten its half-life, that is, the probability of death in a single time period will increase with the generation of antibody, and we assume that the probability of survival of virus in a certain time interval will display a linear downward trend with the generation of antibody;

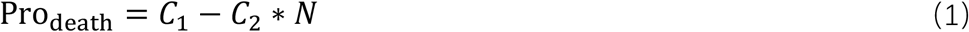

In which *N* represents the time period. *C*_*1*_ *C*_*2*_ is constant.

Assumption 2, we use a random number generator to simulate this stochastic process. We define a replication waiting time. If a virus passes the surviving threshold in assumption 1, that is to say, it does not die in a certain time interval, and its waiting time for replication + time interval > replication period, then this virus will proliferate into two viruses. The replication waiting time of two newly generated offspring viruses = its waiting time for replication + time interval-replication period. If the virus passes the condition of assumption 1, but its waiting time for replication + time period < replication period, then the virus will not replicate, but its replication waiting time = time period + parent virus replication waiting time. A detailed diagram is shown in Figure1.

**Figure1:**
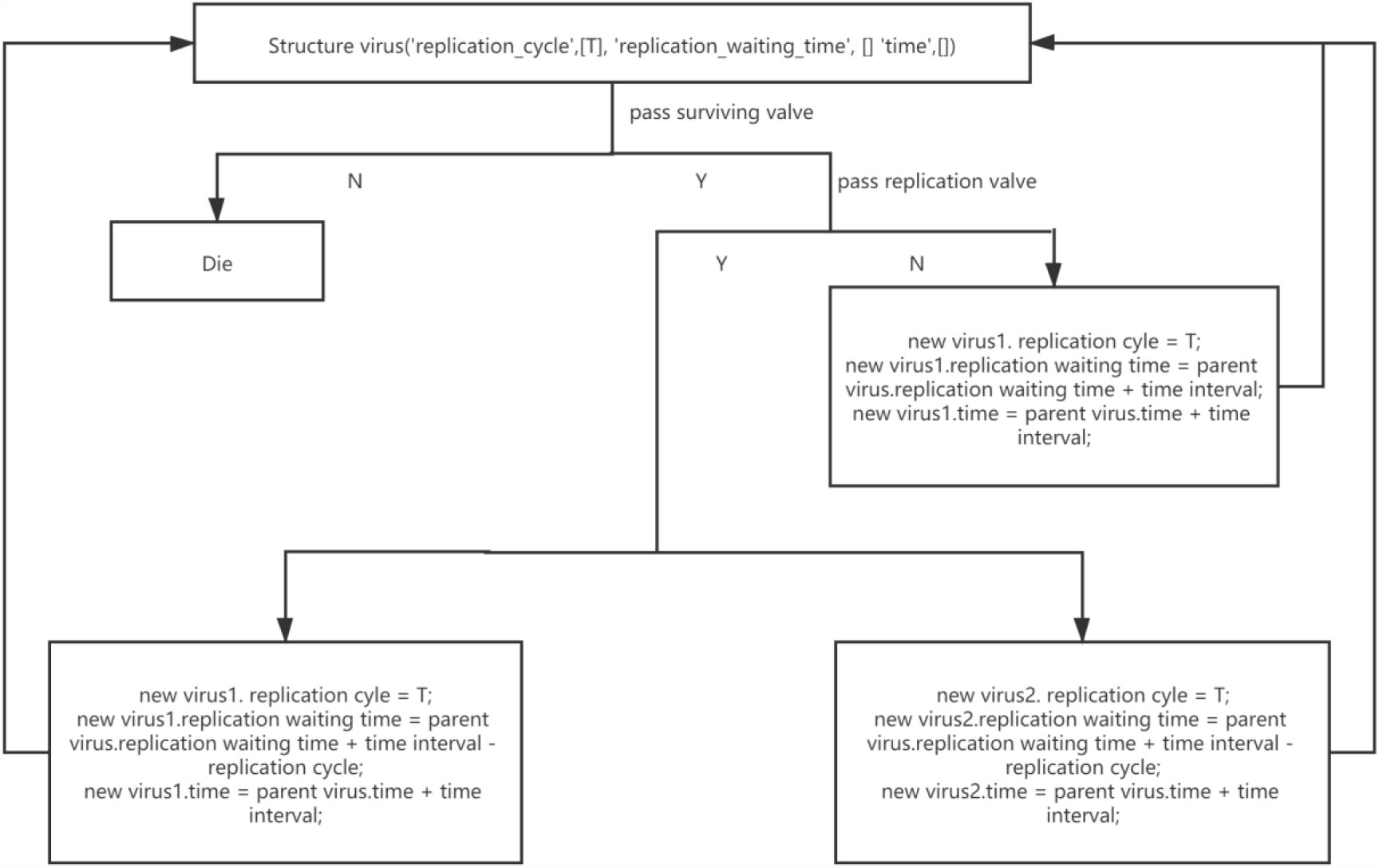
Algorithm workflow illustration.

#### Assumption 3

We assume that for the same virus, the virulence depends on the length of replication cycle. The difference of toxicity of different strains comes from the heterogeneity of sequences, which may be caused the following mutations: mutations on the replication gene directly affects the replication cycle; mutation on other assembly protein genes affect protein assembly efficiency; Mutation in envelope protein genes affects the efficiency of reinfection and transmission, etc. However, regardless of mutations type, its comprehensive long-term effect will eventually lead to the difference of the total virus reproduction efficiency, and this difference can be reflected by the difference of replication cycle. That is to say, for the same virus, the replication cycle of highly toxic strains is shorter and it is easier to cause acute proliferative infection, while that of weaker toxic strains is the opposite.

#### Assumption 4

We adopt a virus accumulation threshold to define infection in our model. We assume that the occurrence of infection requires the peak virus load in host body above a threshold value. If the peak value of the total virus is lower than this threshold, it does not display infection. He or she might be antibody positive but the infectivity is extremely low. If the threshold is exceeded, the infected person will show obvious symptoms of infection, and the larger magnitude passing that threshold, the greater the severity of infection.

According to this model, we simulate the probability of infection caused by different initial doses of strong and weak viruses, and the specific results are shown in the results section.

### 1.2 Define the reproduction number of different virulence strain and its relationship toward host population density

Traditional virus regeneration coefficient R_0_ is defined as the number of susceptible people infected by an infected person after an infection cycle. Here we build a new mathematical model to decompose R_0_ in another way.

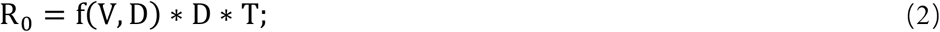

In which D represents the host population density, T represents the infection cycle of virus, and the function f(V,D) represents the probability of infection of virus with toxicity V under the existing population density D. For highly toxic virus, we assume that its infection cycle is shorter, but its f(V,D) is significantly higher than that of low-toxic virus, that is, the probability of infection after single contact is higher than that of low-toxic virus. Different viruses have the same D value in the same social environment. It is worth noting that the function f(V,D) is positively correlated with the toxicity of virus and negatively correlated with the density of host population. In fact, this is easier to explain if we relate this to the social behavior of population animals. Low-density population structure often leads to a longer average time of single contact, while high-density population structure leads to a shorter average time of single contact. For example, if you live in a low population density area, people who come into contact with you will get a longer time during single conversation, while in a big city with a very high population density, more contacts are just within seconds. Actually, transforming (V,D) into the initial virus invasion dose, f(V,D) eventually reflects the dose effect of virus infection. For the same virus strain, the population density d is negatively correlated with the initial virus invasion dose, although it is difficult to express this negative correlation with a formula. Even so, the increase of f(V,D) caused by low host population density can’t make up for the decrease of D caused by the decrease of population density, so the R_0_ value of virus under low host population density is still significantly smaller than the multiplication coefficient R_0_ of virus under high host population density. However, a key point of our virulence evolution theory is that we don’t focus on the absolute value of R_0_ of different strains, but tend to pay more attention to their relative value of R_0_ , that is, the ratio between them. Viral toxicity will evolve towards strains with relatively high R0 value.

## 2 Results

### 2.1 Flaws in the current derivation

A sigmoid relationship between virus dose and its infection possibility is mathematically derived by Jani Anttila et [17]. For details, one can refer to its original paper, here below is the summary of their derivation process:

The pathogen density P is assumed constant for the purposes of this model so that P becomes a parameter in the derived infectivity response. In the following they consider two successive invasions before a susceptible individual becomes infected:

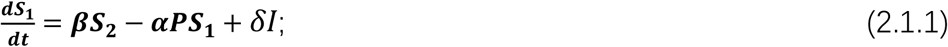

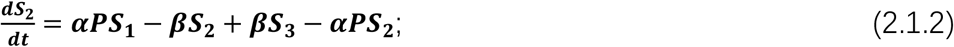

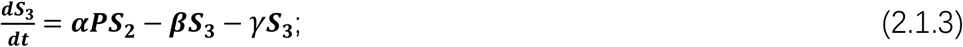

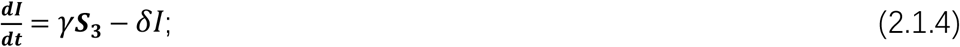

In the above, the contact rate between hosts and pathogens is denoted by α, and the rate at which the immune system kills invading pathogens by β. After two successive invasions the host may become ill with rate γ. Infected hosts, *I*, recover from disease at rate δ. Next, they transformed the above equations into the following ones based on the assumption that the infection and recovery, corresponding to rates γ and δ, is very slow.

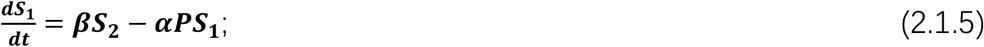

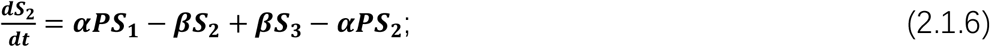

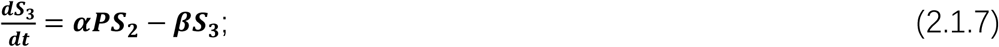

Assuming a quasi-steady state, based on the that the total amount of susceptible hosts stays constant (S_1_ + S_2_ + S_3_ = S), the state S_3_ can be derived as

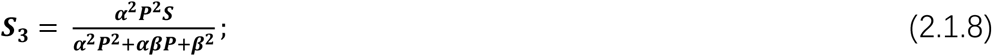

which is a sigmoidal function of *P*. It seems reasonable at first glance, but there are many uncertainties behind this derivation. First, this process does not explicitly stimulate the infection possibility on invading virus dose; Second, in the above group model, the contact rate between hosts and pathogens denoted by α, and the rate at which the immune system kills invading pathogens by β are *P* dependent variables rather than constant. Third, a quasi-steady state assumption further decreases its reliability. Based on these deficiencies, we try to explicitly model the virus proliferation in host using a stochastic model which described in the following part.

### 2.2 Mathematical modeling of stochastic process indicate there is a sigmoidal relationship between virus dose and infection possibility

In this part, virus proliferation dynamics is stimulated using a stochastic model based on the method 1.1. *C*_*1*_ is set to be 0.6, *C*_*2*_ is set to be 0.002, time interval is set to be 10min, the virus replication cycle is set to be 10min. These parameters are not optimized to mimic the actual virus proliferation in host, instead, those number are selected with more consideration on computational efficiency. An up-regulation of *C*_*1*_ and down-regulation of *C*_*2*_ will lead to a faster proliferation process and generate more viruses at different time points. This will lead to a more reliable stimulation with a higher computation cost. On the other hand, the opposite direction will speed up the stimulation at the cost of accuracy. We set these values based on a balance between computational reliability and efficiency. The initial virus dose was set from 1 to 50. 5000 iterations were enforced for the simulation.

Maximal virus loading distribution at different invading dose is shown in figure2A. It can be noticed that the virus peak quantity in host is positively positive related with the invading doses, but it is not a linear relationship. The relationship between infection possibility and initial virus dose is shown in figure2B with different infection threshold.

**Figure2A:**
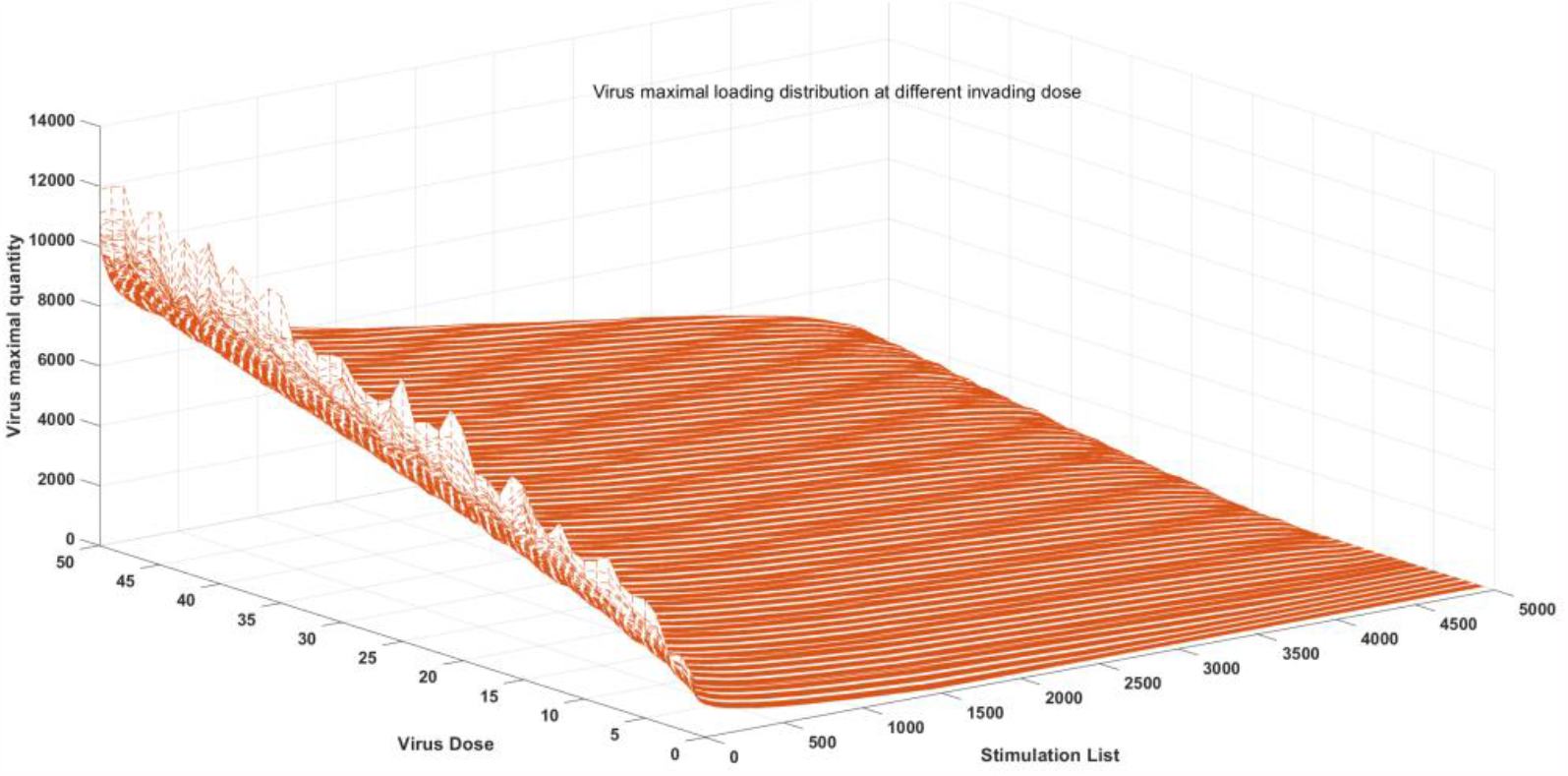
maximal virus loading distribution at different invading dose

As displayed in fig.2B, we can see there is a sigmoid relationship between infection possibility and initial virus dose. It first surprised us since the modeling results indicated it was not a logistic curve. Then we realized the accumulation effects on virus infection. This accumulation effect makes it does not follow the simple principle of probability calculation. For instance, if the infection probability of single virus infection is *P*, according to simple probability calculation, the infection probability of N viruses should be 1-(1-*P*)^N^, then the relationship should turn out to be a function with a persistent declining derivative. However, because the virus proliferation quantity is not a discontinuous one, the accumulation effect makes the infection curve sigmoidal rather than exponential. This sigmoidal relationship is validated by many wet experiment evidences and is important for our prediction of virulence evolution described in part 2.4.

**Figure2B:**
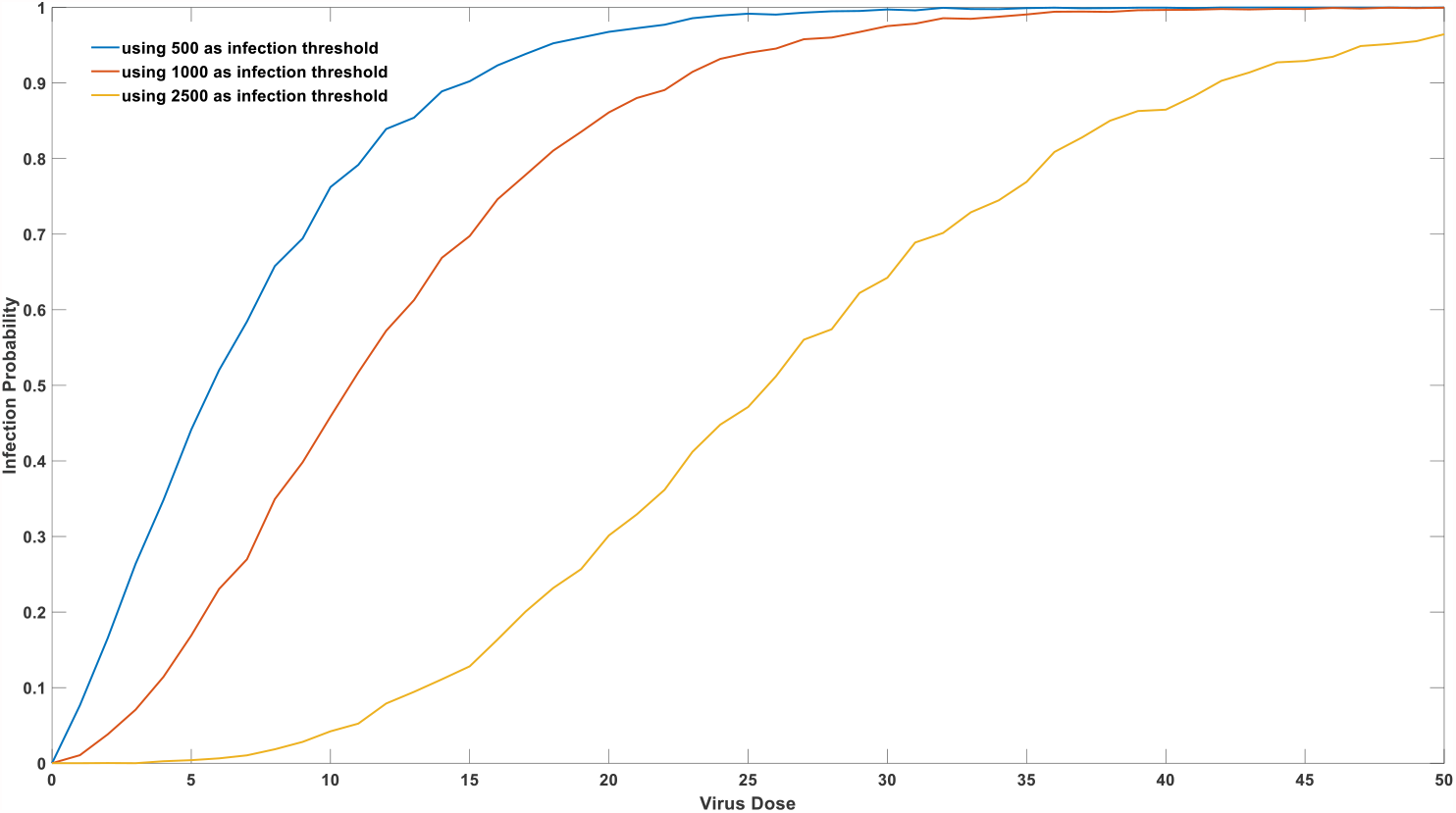
relationship between infection possibility and initial virus dose using different infection threshold.

### 2.3 Three factors influence the infection severity: viral virulence, host immunity and virus entry dose

It has been empirically noticed long time that the individual viral virulence played an important role in the infection severity [18-20]. It can be seen from the Fig.3A, for people with same immunity with same d value, in the random simulation of 5000 infection cases, the maximum host viral load of strong virulence virus is obviously larger than that infected by weak virulence virus. This could mathematically explain the influence of COVID-19 virulence in mortality rate of SARS-CoV-2 infection.

**Figure 3A:**
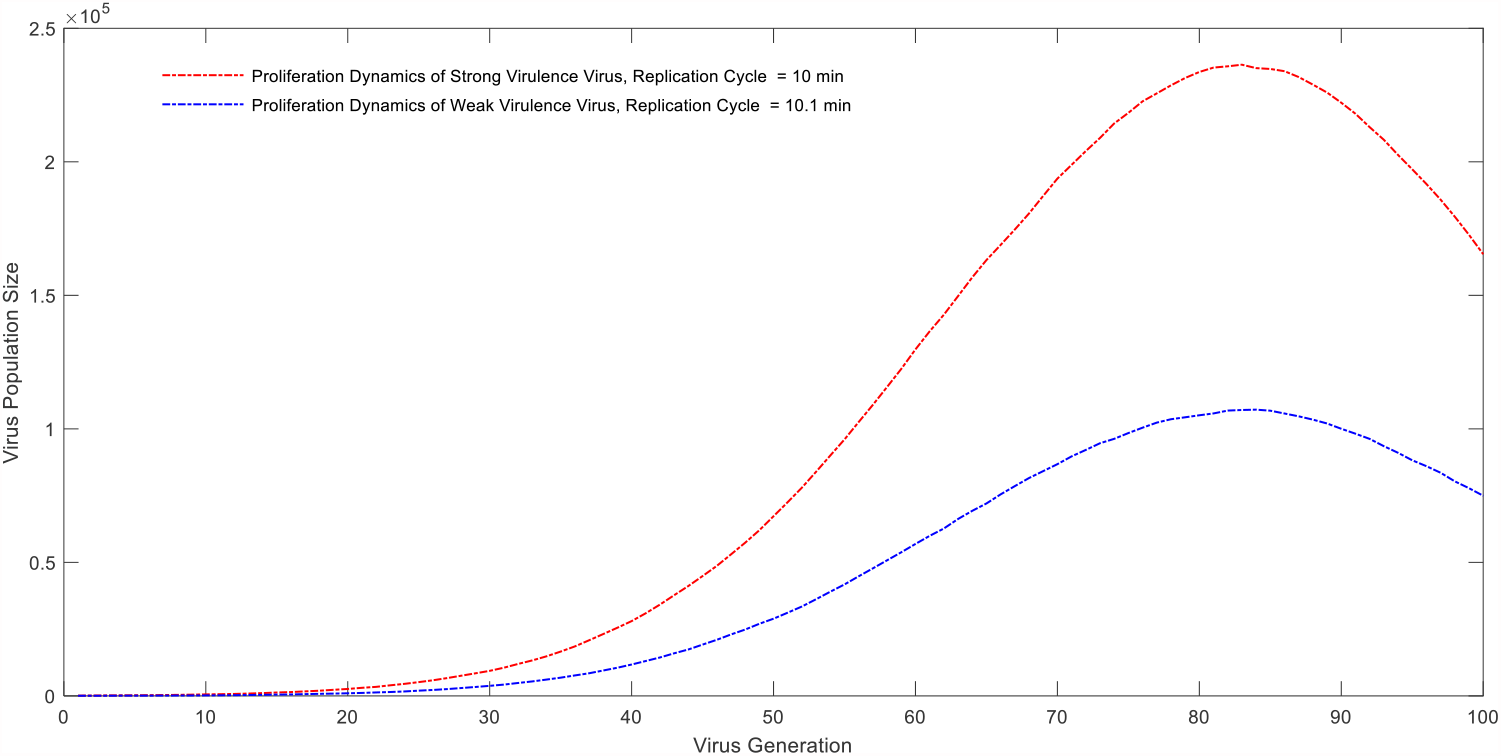
Proliferation dynamics of different virulence strains in host body

As far as individuals are concerned, people with strong immunity are less likely to be infected with viruses than people with weak immunity, and the mortality rate of virus infection is significantly lower than that of people with weak immunity. As shown in Figure 3B, yellow curve represents the virus number fluctuation in the host with strong immunity. Green color represents the virus dynamic in host body with medium immunity. Blue curve represents the proliferation dynamic in host body with weak immunity. From this figure, we can see the virus quantitative change is closed related to the host immunity, strong immunity will inhibit virus in-vivo proliferation.

**Figure 3B:**
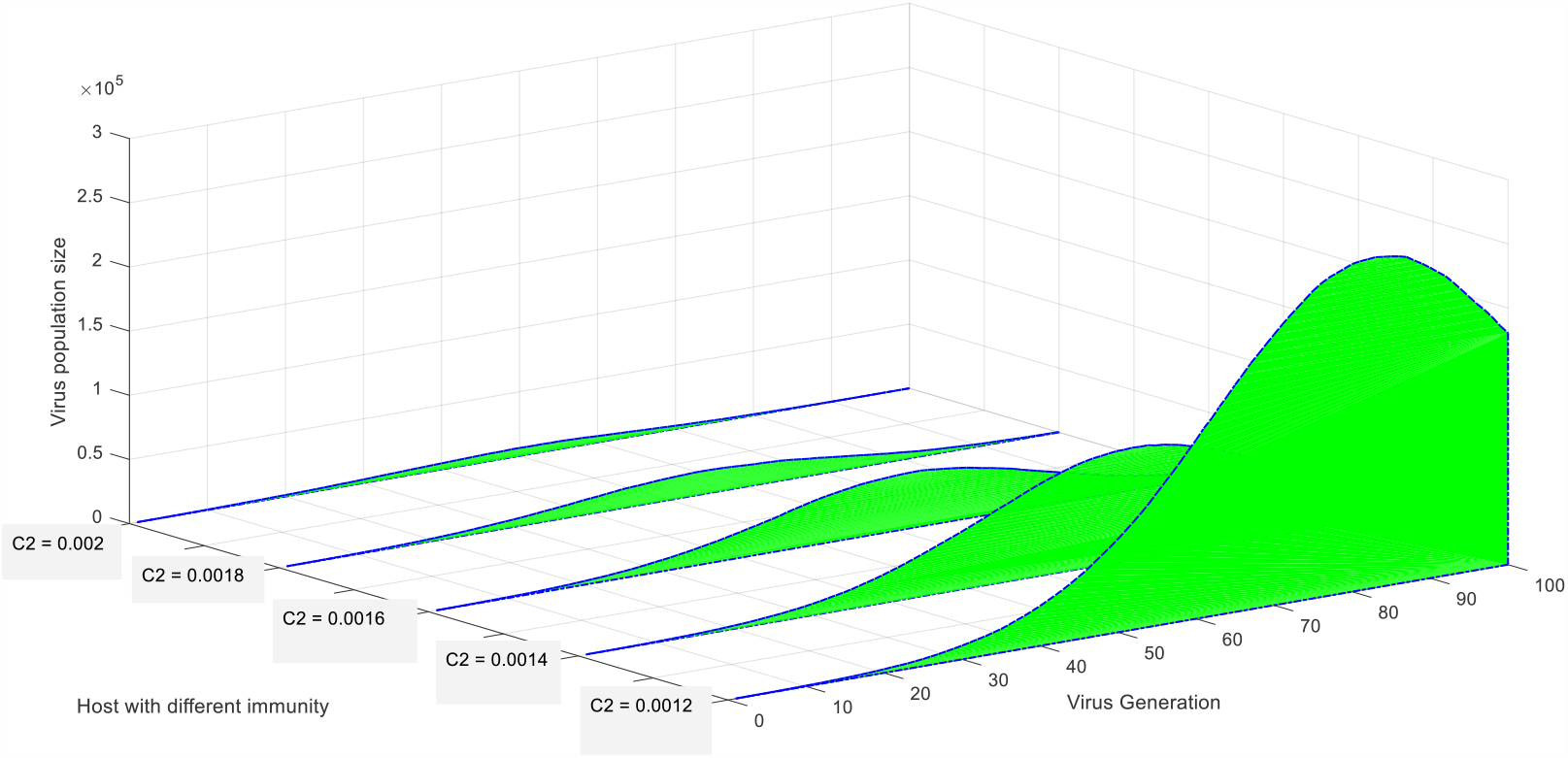
Virus proliferation dynamics in host body with different immunity

However, when the virus concentration in the environment rises to a certain degree, the number of inhaled viruses will increase proportionally, all people will lose their barrier function against viruses, and infections will be induced unavoidably. However, people with strong immunity are not easy to develop into symptomatic and severe cases as shown in Figure 3C.

**Figure 3C:**
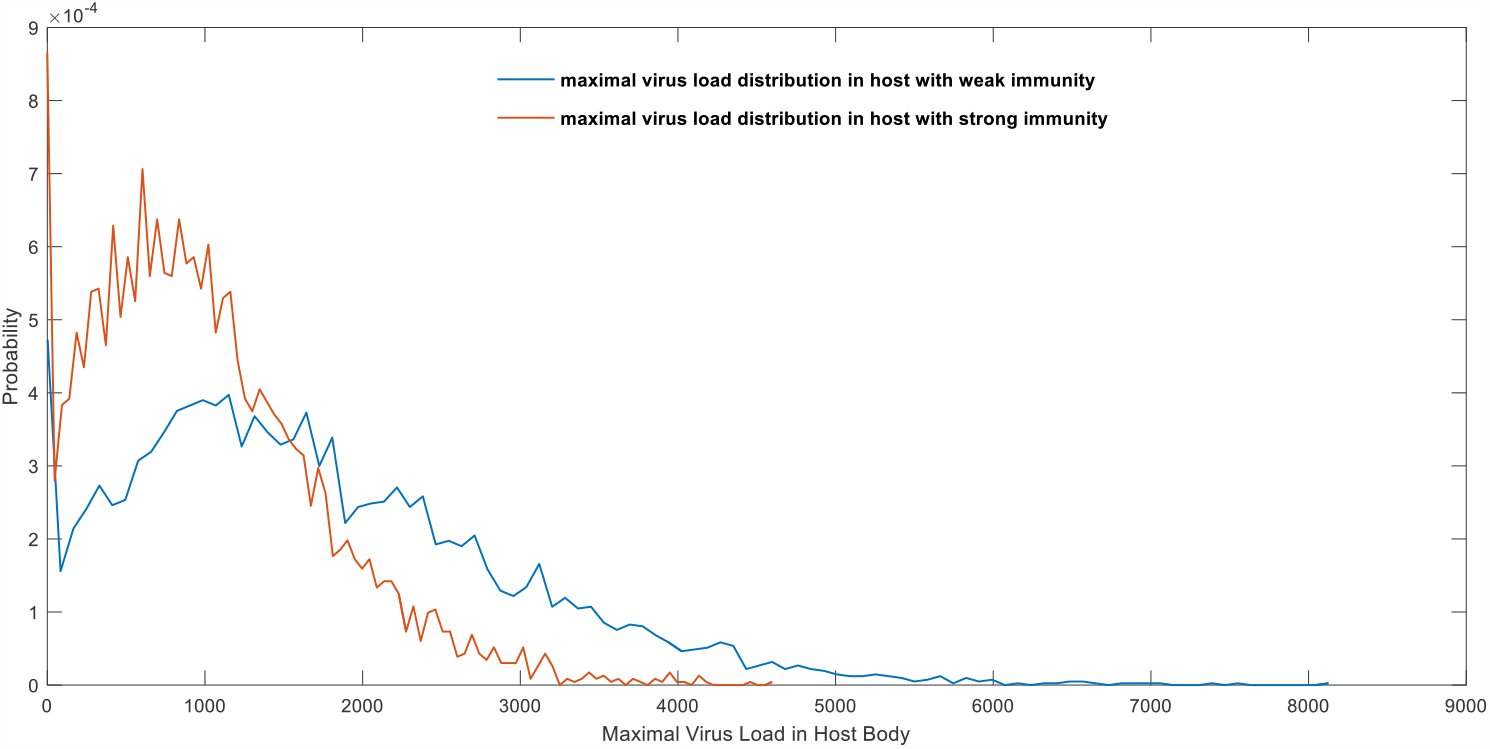
Virus peak quantity distribution among individuals with different immunity

In our model, immunity is represented as c2. We assume that c1 is the same for every individual while c2 varies among people with different immunity. People with strong immunity such as young people will have a quick immune response against the invading virus and produce larger amount of antibody to fight against it. Therefore, parameter c2 in people with strong immunity is bigger than c2 in weak immunity people. It means the virus’s half-life in strong immunity hosts will be shorter than those in weak ones throughout infection cycle. It can be seen from the Fig. 3C, for people with strong immunity, 5000 random simulations of virus invading cases indicate the maximum viral load in the host body is obviously smaller than that of people with weak immunity. The proportion of individuals with strong immunity who eventually develop into individuals with high viral load is very small, while the proportion of individuals with low immunity who eventually develop into individuals with high viral load is significantly higher than those with strong immunity. This can explain the significant age difference in mortality rate of SARS-CoV-2 infection.

It also been noticed the average infection age is declining during the epidemic which is hard to explain [21]. It can be seen from Fig. 3D that when people with the same immunity are invaded by virus with strong virulence in a small dose (n=2), 5,000 random simulation cases show that the 45.48% of individuals encounter with infection (using 100 as the infection threshold). However, proportion of samples (26.7%) still show the characteristics of high virus load (using 500 as a severe infection threshold), as displayed in the blue line of Fig. 3D. When invaded by virus with a weaker virulence in a large dose (n=50), the random simulation of 5,000 samples show that a larger proportion of susceptible population (72.14%) suffer infection, but less samples display the characteristics of high viral load (3.3%), as shown in the red part of Fig. 3D. This can well explain the average infections age is decreasing in the later period of the epidemic. At the very beginning of the epidemic, virus is highly toxicity, but the concentration in the environment is low, and the invading virus dose is relatively small. For young people with strong immunity, the peak load of most viruses cannot overcome the infection threshold. Although infection is less likely to occur, once infection happens, compared with those infected in the later period of the epidemic, there is a greater risk of turning into severe condition or even death. To the contrast, the virus toxicity is weak in the later epidemic period. However, a large amount of shedding viruses has accumulated in the environment, and the number of invading viruses has dramatically increased. The increase of the number of invading viruses will make the peak viral load in host pass the threshold of infection. Therefore, young people would lose their shielding effect on SARS-CoV-2, and the epidemic situation is characterized by youthfulness. However, due to the reduced toxicity of the virus, the peak viral load in infected people often does not reach the valve of severe illness or death, so the mortality rate and severe illness rate in the later period are obviously reduced.

**Figure 3D:**
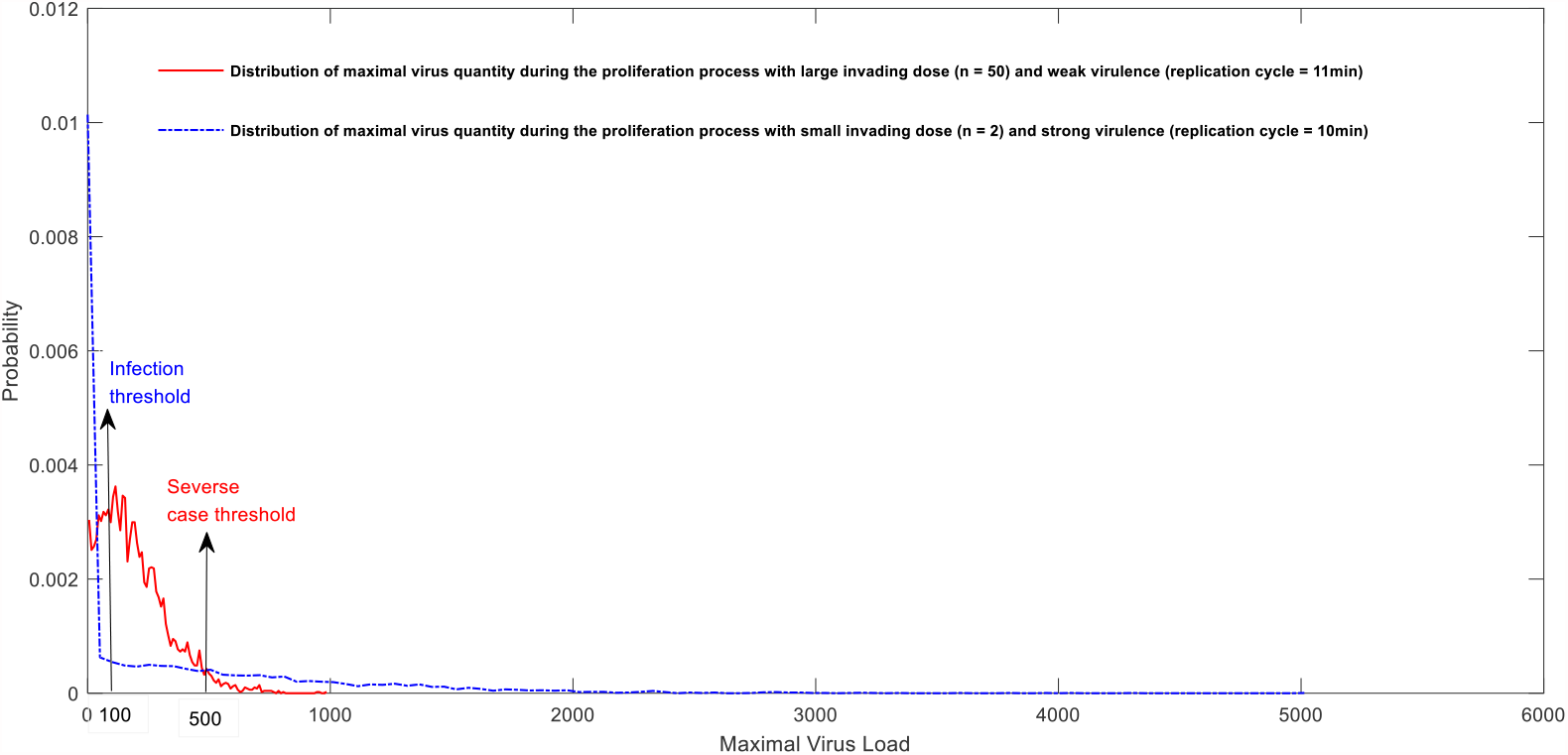
Modeling of individual infection with different virus invading dose and virulence.

### 2.4 Population effect on virus virulence evolution

Real-time statistical mortality rate proves that the toxicity of viruses is constantly changing, and different countries and regions have different trends. For example, the toxicity of SARS-CoV-2 in India is relatively low, while some Latin American countries such as Mexico is extremely high. For the United States, the mortality rate experienced a rapid decline in the early stage and fluctuations in the later stage. From a global perspective, the mortality rate of COVID-19 infection also experienced a rapid decline in the early stage and a constant fluctuation in the later stage. If the data are authentic and credible enough, this slight fluctuation of late mortality is statistically significant, which indicates that the virulence of SARS-CoV-2 is evolving all the time. The relationship between host contact density and virus toxicity well follow the principles of ecological balance and ecosystem evolution. The research on the evolution of virus toxicity is an old research topic. According to the traditional view, there is a subtle trade-off relationship between virus transmission coefficient and virus toxicity [12]. Viruses with extremely high toxicity and extremely low toxicity will cause less transmission potential. The best toxicity is the virus strain that can bring the maximum transmission coefficient. Many studies also point out that this best toxicity is not fixed, and is related to the characteristics of the virus itself and the interaction with the host. However, there is no mathematical model that can successfully explain the relationship between virus toxicity and host population contact density. We try to establish such a model to better analyze and explain the relationship between virus virulence evolution and host population contact density.

The establishment of this model is based on the virus microscopic proliferation model. The details of this model are described in materials and methods section. We assume that two viruses with different toxicity have the same R0 under the condition of initial population density. The differences between these two strains are V and T. We assume that the highly toxic strain

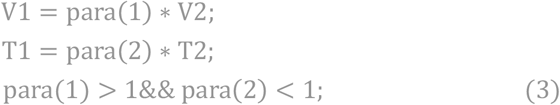

Since they have same R_0_ ,

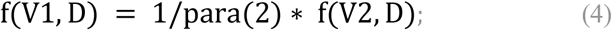

So when D changes, how does this balance break? If we think that f(V, D) is linearly related to V*Time_contact, that is, the probability of single infection is linearly related to the total amount of virus inhaled once, then this balance will not change at different D value. However, the actual situation is not the case. According to our simulation, the sigmoid relationship between the infection probability and the virus entry dose is shown in Fig.4A.

**Figure4 A:**
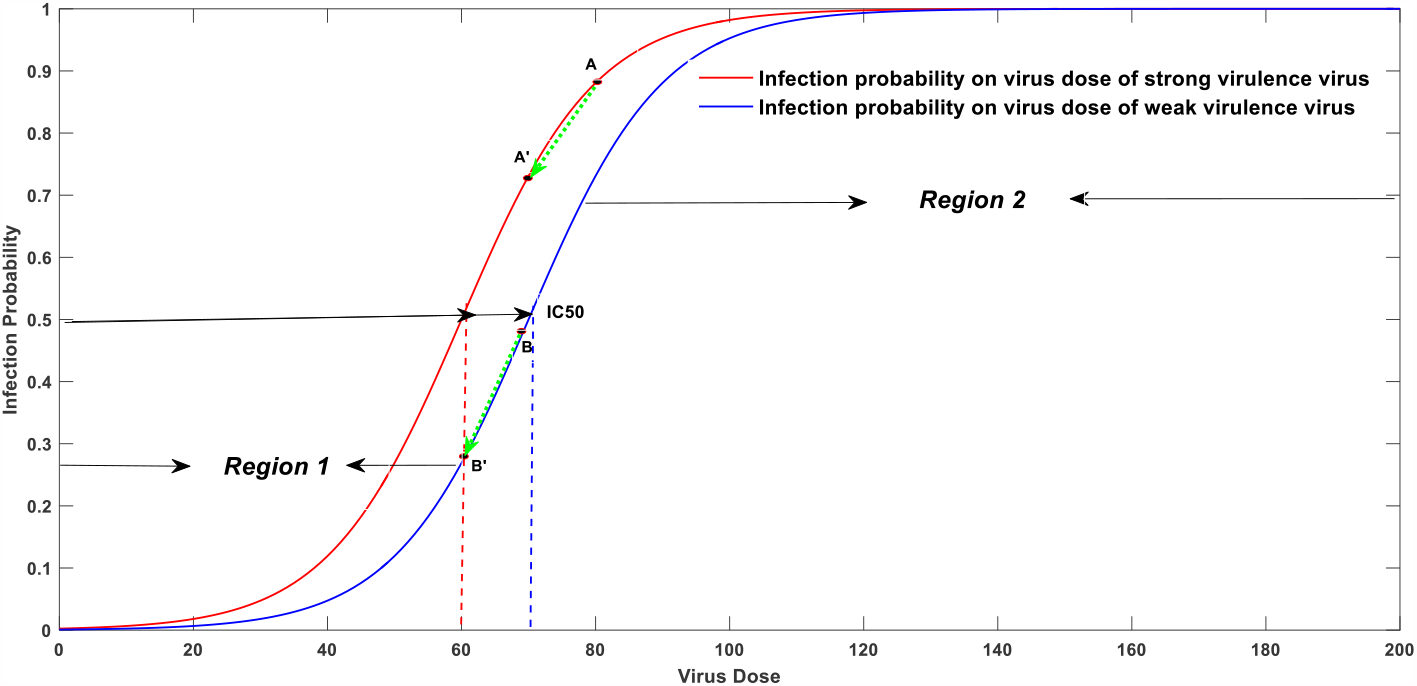
Mathematic representation of infection probability on invading dose with two different virulence strain.

It can be mathematically represented as

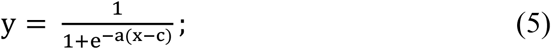

As shown in Fig.4A, c value in weak virulence virus is bigger compared to strong virulence virus. Parameter a is set to be 0.1 for both strains, c is set to be 60 for the strong virulence strain and 70 for the relatively weak one. When population density increase, the problem turns out to be which strain has a relatively larger R0. As shown in Fig.4A, the original average inhaled virus dose for strong virus is marked as point A. The new point after increasing population density is marked as point A’. The original average inhaled virus dose for weak virus is marked as point B and the new point after increasing population density is marked as point B’. Since the change of population density will decrease the average inhaled virus doses with the same magnitude for both strains,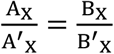.Then the problem turned out to be 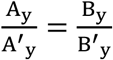 ?. If 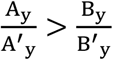,strong virulence virus will engage a bigger R_0_ declination compared to the weak one, virus will evolve into the weak direction. It is the opposite direction otherwise.

It is difficult to mathematically derive the solution for us to demonstrate which strain will have a bigger R_0_ value with increased host population. Therefore, a computational modeling approach is used to infer which strain will dominate the evolution.

We perform the stimulation using different parameter, the simulation results are displayed in figure4B. Population density change magnitude represents the step size caused by increasing population density which is same as 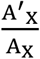 in figure 4A. This variable was set from 0.5 to 0.95 with a 0.05 interval. Z coordinates is represented as 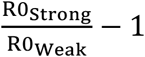. When it is positive, the strong virulence strain has the advantage over the weak one. X coordinates represent the virus dose which is the same as figure4A and it has an opposite trend against population density. From figure 4B, we can infer that at the relatively dense population conditions, as shown in the curve with x value range from 0 to 60, a further increase in population density will do more favor to the evolution of weak virulence virus. 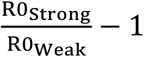 is a negative number which means environment is beneficial in selecting the weaker virulence strain. However, at relatively sparse population density, a raise in the population density will enable strong virulence virus an evolution advantage, as shown in the curve with x value range from 80 to 200. This is a very interesting finding, it seems that population will protect themselves only beyond certain threshold. Below this threshold, population growth will harm the specie, at least in the perspective of guiding virus evolution. That means the Allee-effect theory in ecology is conditional when we consider the population effect against infections.

This mathematic modeling indicates before reaching the IC50 turning point, declination of virus invading doses caused by increase population density will attenuate the overall virulence. However, the trend is opposite when it passes the turning point which means under very sparse population density level, the increase of population density will reinforce the virulence. So how to judge which region are we now at? According to mortality data, we noticed that the high-density countries such as India, Singapore, Japan, South Korea, Philippines and so on, have a relatively low mortality rate compared to the rest world. Therefore, we predict the average inhaled dose per contact is below IC50 at most countries during COVID-19 infection. The majorities of the world locate in the first area depicted in Fig 4A.

**Figure4B:**
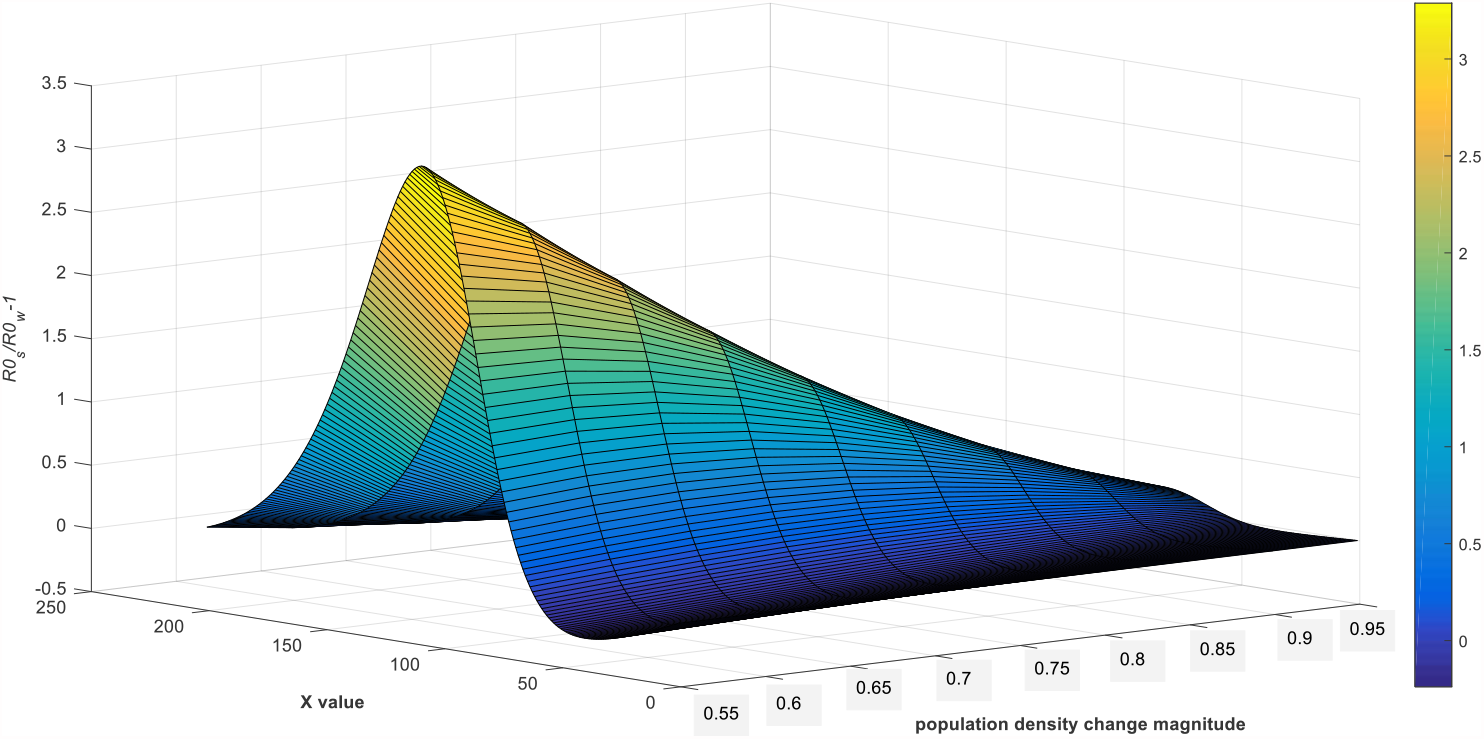
Numerical diagram of relative ratio of R_0_ values of two different virulence strain.

As shown in Fig.4A, c value in weak virulence virus is smaller compared to strong virulence virus. It is very difficult to mathematically demonstrate which strain will have a bigger R0 value with increased host population. Therefore, a computational modeling approach is used to infer which strain will dominate the evolution.

We perform the stimulation using different parameter, the simulation results are displayed in figure4B. From figure 4B, we can infer that at the relatively dense population conditions, a further increase in population density will do more favor to the evolution of weak virulence virus. However, at relatively sparse population density, a raise in the population density will enable strong virulence virus an evolution advantage. This is a very interesting finding, it seems that population will protect themselves only beyond certain threshold. Below this threshold, population growth will harm the specie, at least in the perspective of guiding virus evolution. That means the Allee-effect theory in ecology is conditional when we consider the population effect against infections.

Our finding can give a better explanation on the mortality difference among different countries. In a long time range, the mortality in densely populated area is significantly lower than that in moderately populated area. The highest mortality records always come from countries with medium population density. This recovery is not our most critical point yet. Since we notice the sensitive relationship between virus virulence and host population, what can we do to perform some manual intervention to promote the virulence attenuation? We proposed that the protection measures such as face mask and social distance might be more efficient than vaccination, especially in long term. Wearing a mask cannot completely isolate the virus, but it can effectively reduce the total virus invading dose, thus reducing the infection probability of the wearer. At the same time, as shown in Fig. 2B, the peak viral load of a single infection is also positively correlated with the initial viral invasion, so wearing a mask can not only reduce the probability of infection of the wearer, but also effectively reduce the severity of the infection which has been reported in many literatures [22-25]. Meanwhile, and maybe the most important viewpoint that people always ignored, face mask and social distance will accelerate virus evolution towards low toxicity. It will lead to less virus entry dose which is the same effect as increasing the population. Within the blue lines depicted in figure 4C, the decrease of average virus entry dose will promote the virus evolution toward low toxicity. While it is difficult to mathematically evaluate its driving force, a real case is astonishing. This extreme and interesting phenomenon is that under the extremely strict epidemic prevention measures in Wuhan, there were more than 300 infections in the later period of the epidemic and they were all asymptomatic, indicating a very rapid virulence declination within 3 months strict lockdown policy [26]. When we talk about protection measures and social distances, it always comes first that these policies can directly decrease the infection possibility. Very few people realized its power in attenuating virus virulence in long term. Its function in virulence attenuation might even be more significant and valuable compared to its explicit effect on reducing infection cases. Because no matter how strict the policy is, it is impossible to complete block the transmission. However, strict epidemic prevention measures have the capacity to direct the virus evolve into the weaker strain before a complete elimination within several month.

Interestingly, herd immunization will not affect the value of f(V, D), because the group immunization process will not affect people’s social behavior, that is, the single contact time will not increase, which is different from the effect of lowering population density. However, it could enable people with alleviated symptom thus can reduce the total amount of environmental virus released from infected cases. As shown in the blue section, reduced virus dose will promote the virus evolve into the weak direction. Therefore, large scale vaccination is not only able to explicitly reduce R0, but can also accelerate overall virulence attenuation.

## 3 Discussion

In this paper, we proposed a modeling approach to stimulate the virus proliferation dynamic in vivo. Our model is based on a simple logic: survive to replicate. This stochastic stimulation indicate there is a sigmoid relationship between inoculum dose and infection possibility. The stochastic modeling underpinned the mechanism of virus dose effect. What’s more, it provides a new idea in the future vaccine design and vaccination strategy optimization. Since the vaccine is designed not to reproduce itself, the model will turn out to be: survive or die. Using a stochastic modeling approach, one can stimulate the virus quantity change through time. By setting a criterion toward antibody generation, one can estimate the protection efficiency per injection. This criterion could be the overall timespan above certain quantity threshold. By doing so, instead of waiting the clinical feedback, we can mathematically optimize the inoculum dose, including the vaccination strategy.

Based on this sigmoid correlation, many strange events and phenomenon during COVID-19 epidemic can be explained. For example, why the new infected people are tending to be younger, indicating individual with strong immunity are losing their protection shield against the virus. Besides that, we found that there is a sensitive relationship between the changes of SARS-CoV-2 toxicity and invading virus dose per contact. High population density will naturally reduce the interaction time per contact. This will lead to less virus dose per contact which will further promote the virus evolve into the less fatal one in long term. Social distance and protection measures will artificially reduce the virus dose per contact which could also help the virus virulence attenuate to a mild one. At a time when the majority of media and publics pay extremely high expectation on vaccine, it is hard to deny the classic herd immunity theory. However, we have to be cautious since contradictions have emerged, both on practice and theoretical level. It is a time that we should redefine “herd immunization”. Herd immunization should not stand for a quick virus elimination after high vaccination coverage. It will lead to a long-virulence decaying process after vaccination which is already proved by a low morality in the resurged epidemics in some well-vaccinated country such as Britain and Israel. we cannot completely eliminate the virus by group immunization but we can tame them into a mild one.

Furthermore, when we review the success of Chinese anti-epidemic efforts, instead of focusing on their explicit effects, we should realize the persistent virulence-attenuation effects on these simple quarantine strategies. A theory is unreliable if it does not match with reality, meanwhile, a phenomenon might be coincidence without physical theory support. As the case of COVID-19 virulence evolution, our virus virulence evolution theory is consistent well with the existing statistical evidences. Therefore, if we cannot eliminate the viruses, should we tame them?

## Data Availability

I have followed all appropriate research reporting guidelines and uploaded the relevant EQUATOR Network research reporting checklist(s) and other pertinent material as supplementary files, if applicable.

## Acknowledgement

We thank Dongqing Wei from Shanghai Jiao Tong University for fruitful initial discussions, Yushan Zhu from Tsinghua University, Jian Song, Peiyan Guan, XiangYong Li, and Zhenlin Wei from Dezhou University for helpful conversations, comments, and clarifications. This research is funded by Dezhou University.

## Supplementary materials

Matlab codes can be accessed at:

***https://github.com/zhaobinxu23/Mathematics_underpinning_the_dose_effect_of_virus_infection_and_its_application_on_covid-19_virulence_evolution***

